# Influence of BMI-associated genetic variants and metabolic risk factors on weight loss with semaglutide: a longitudinal clinico-genomic cohort study

**DOI:** 10.1101/2024.10.31.24316494

**Authors:** Matthew E. Levy, Natalie Telis, Kelly M. Schiabor Barrett, Alexandre Bolze, Douglas Stoller, Christopher N. Chapman, C. Anwar A. Chahal, Daniel P. Judge, Douglas A. Olson, Joseph J. Grzymski, Nicole L. Washington, William Lee, Elizabeth T. Cirulli

**Author notes:** Corresponding author: Matthew Levy, PhD. Helix. Mail: 101 S. Ellsworth Avenue, Suite 350, San Mateo, CA 94401.

## Abstract

**Background:** Individual weight loss response to the GLP-1 receptor agonist semaglutide varies considerably, with many possible contributing factors. Leveraging multiple clinico-genomic cohorts, we analyzed differences in weight loss trajectories according to patient characteristics, including a polygenic score (PGS) and metabolic risk factors, in semaglutide initiators with BMI ≥27 kg/m^2^.

**Methods:** This longitudinal study utilized clinical-grade exome sequencing and electronic health record data from six U.S. cohorts within the Helix Research Network (n=134,806). A BMI PGS was calculated using 26,941 variants. Twelve-month weight loss trajectories were modeled using mixed effects models, and associations with demographics, PGS, comorbidities, medications, and laboratory results were evaluated.

**Findings:** Among 1,923 semaglutide users, the mean pretreatment BMI was 38.4 kg/m^2^. For those on doses ≥1.7 mg, the mean body weight reduction was 7.3% at 6 months and 9.9% at 12 months. Over 12 months, low PGS was associated with an adjusted 1.5% and 1.8% additional weight loss compared to intermediate and high PGS, respectively (both p<0.01). Male sex, type 2 diabetes, hypertension, obstructive sleep apnea, and non-alcoholic fatty liver disease were each associated with 1.2%-1.9% less weight loss (all p<0.05). In type 2 diabetes, each 1%-increase in pretreatment hemoglobin A1c was associated with 0.6% less weight loss (p=0.0019).

**Interpretation:** Among adults with overweight or obesity, a lower genetic predisposition to obesity is linked to greater weight loss on semaglutide. Additionally, metabolic health significantly impacts the drug’s effectiveness. These findings underscore the importance of precision medicine in obesity management.

**Funding:** Renown Health Foundation. Nevada Governor’s Office of Economic Development. HealthPartners.

## Introduction

The emergence of glucagon-like peptide 1 receptor agonists (GLP-1 RAs) for weight management have introduced a promising new treatment option for the high global prevalence of overweight and obesity. The GLP-1 RA semaglutide has been approved by the US Food and Drug Administration and European Medicines Agency for both type 2 diabetes (T2D) management (branded as Ozempic) and weight loss (branded as Wegovy). Multiple phase 3 randomized controlled trials have demonstrated its efficacy for weight loss.^1–6^

However, there has been considerable heterogeneity in weight loss outcomes among users. In the STEP 1 trial, weight loss among 2.4-mg semaglutide users exhibited a Gaussian distribution, with 14% failing to achieve ≥5% weight loss alongside a remarkable 33% who achieved weight reductions of ≥20% by 68 weeks.^1^ Real-world effectiveness studies also show variability, with 13% to 60% of users failing to achieve ≥5% weight loss after 6 or 12 months.^7–10^

While several factors are known to influence the amount of weight lost with semaglutide, most of the variation remains unexplained. Studies show that females, on average, experience more weight loss than males,^8,10,11^ and individuals without T2D lose more weight than those with T2D,^1,2,7,8,10^ though some of this disparity may be due to dose differences. The extent to which other patient characteristics–including demographic factors, genetic variation, other comorbidities, concurrent medication use, and clinical and laboratory indicators of metabolic health–impact individual responses to semaglutide remains unclear.

Leveraging the Helix Research Network,^12^ a network of US-based clinico-genomic cohorts, we aimed to identify demographic, genetic, pharmacologic, and metabolic factors associated with weight loss trajectories over 12 months following the initiation of subcutaneous semaglutide in adults with overweight or obesity. This is one of the first large-scale studies^13–16^ to incorporate a polygenic score (PGS; an aggregate of contributions from multiple genetic variants^17,18^) in an evaluation of treatment response, and the first, to our knowledge, for a weight loss medication. This investigation is an important step toward refining and personalizing GLP-1 RA-based weight loss expectations and strategies.

## Methods

### Study design and participants

In this observational longitudinal clinico-genomic study, we retrospectively collected electronic health record (EHR) data from adult patients enrolled across six Helix Research Network study protocols open to the general patient populations at different U.S. healthcare organizations. The participating protocols are DNA Answers (St. Luke’s University Health Network, Pennsylvania), the Genetic Insights Project (Nebraska Medicine), the Healthy Nevada Project (Renown Health, Nevada), In Our DNA SC (Medical University of South Carolina), myGenetics (HealthPartners, Minnesota), and The Gene Health Project (WellSpan Health, Pennsylvania). EHR data were transformed into the Observational Medical Outcomes Partnership (OMOP) Common Data

Model (CDM) version 5.4, encompassing a median of 10.4 years of EHRs per patient (interquartile range [IQR]: 4.9-18.3). Additionally, saliva samples were collected from participants and underwent Exome+^®^ sequencing at Helix between Feb 2018 and Jun 2024, as previously described.^19,20^ Study protocols were reviewed and approved by their respective Institutional Review Boards (projects 956068-12 and 21143). All participants provided written informed consent prior to participation, and all data used for research were deidentified.

For this analysis, individuals with linked EHR and Exome+^®^ sequencing data were included if they met the following criteria: 1) having ≥2 prescriptions for subcutaneous semaglutide spanning ≥3 months in the first year, signifying likely ongoing use for a minimum 3-month duration; 2) BMI ≥27 kg/m^2^, consistent with weight loss treatment guidelines; 3) absence of other medications, procedures, or conditions expected to influence weight, including tirzepatide prescription(s) in the prior year, a history of bariatric surgery, malignancy in the prior year, or pregnancy in the prior nine months; and 4) having ≥2 on-treatment follow-up weight measurements taken ≥2 months apart, spanning 3-12 months of treatment. The OMOP CDM concept sets used to define inclusion criteria and other measures are available as a supplemental data file.

### Treatment

Intervals of subcutaneous semaglutide treatment were defined using ≥2 prescription dates for Ozempic or Wegovy (both Novo Nordisk). To comprehensively characterize real-world weight loss patterns among all semaglutide users, including those with or without T2D, individuals were included regardless of whether they were prescribed Ozempic (labeled for T2D) or Wegovy (labeled for weight loss), and regardless of their dosing patterns. To maximize specificity in identifying patients with prescription patterns indicating ongoing subcutaneous semaglutide use for ≥3 months, patients were considered treated if they had multiple prescriptions covering at least a 3-month period within the first year. Prescription fulfillment and compliance data were not available. The semaglutide initiation date was defined as the date of first prescription, while the estimated end date was set as three months after the last documented prescription, as stop dates were not consistently available. The dose of semaglutide was treated as time-varying and classified as 0.5 mg, 1.0 mg, 1.7 mg, 2.0 mg, or 2.4 mg at each time point using the most recent prior prescription. In instances with multiple doses prescribed on the same date, dosing intervals were interpolated based on the expected dose escalation schedule. Further information about prescription records and data preparation are available in the appendix (pp 1-2).

### Outcome

Among all body weight measurements recorded during healthcare visits up to 12 months after starting semaglutide, percentage weight change was calculated as follow-up weight minus pretreatment weight, divided by pretreatment weight. The pretreatment weight was the last weight measurement obtained on or before the treatment initiation date, within a maximum interval of six months. If patients newly had a prescription for a different GLP-1 RA or tirzepatide, a bariatric surgery procedure, a malignancy diagnosis, or pregnancy during the follow-up period, weight measurements were censored at those dates. Data quality control procedures were implemented to ensure the validity of weight measurements, including checks for implausible changes, which are detailed in the appendix (p 3).

### Covariates

The Exome+^®^ data from each participant were used to calculate a BMI PGS and determine genetic similarity group compared to the Phase 3 data of the 1000 Genomes project using the Admixture software as previously described.^21^ The BMI PGS (PGS001228) incorporated 26,941 variants and was computed based on genotype probabilities.^22,23^ Among 96,914 Helix Research Network participants with no documented history of GLP-1 RAs or other anti-obesity medications, the PGS showed a positive correlation with their most recent BMI (R-squared=0.082, p<0.001), demonstrating its validity in representing genetic predisposition toward higher BMI in this study population (appendix, p 4). Percentiles were calculated within each genetic similarity group, as previously described.^24^ The PGS was then classified into low (bottom quintile), intermediate (quintiles 2-4), and high (top quintile) categories, which were similarly associated with BMI (appendix, p 5). This categorization scheme created distinct groups for low and high genetic risk while maintaining adequate sample sizes in each category.

Other covariates were derived from EHR data. Demographic characteristics included health system, age, sex, and EHR-recorded race and ethnicity. Comorbidities were defined using diagnosis codes and included T2D, hypertension, hyperlipidemia, gastroesophageal reflux disease (GERD), obstructive sleep apnea, and non-alcoholic fatty liver disease (NAFLD).

Recent use of a different GLP-1 RA (albiglutide, dulaglutide, exenatide, liraglutide, or lixisenatide) or other anti-obesity agent (orlistat, naltrexone-bupropion, phentermine, phentermine-topiramate, or setmelanotide) were defined by the presence of prescription(s) in the prior year. Concurrent medication use was determined based on prescriptions for other anti-obesity agents, insulin, metformin, antihypertensive agents, or lipid-lowering agents during the same period as semaglutide. Pretreatment clinical and laboratory test results, including blood pressure, hemoglobin A1c, and serum lipids, were defined using the most recent measurements available prior to semaglutide initiation.

### Statistical analysis

Patient characteristics were summarized both overall and stratified by the highest semaglutide dose reached. Percentage changes in body weight were summarized at 3, 6, 9 and 12 months post-initiation using measurements within a window of ±30 days.

Using all on-treatment follow-up weight measurements, linear mixed effects models with linear and quadratic terms for time were employed to model percentage weight change through up to 12 months. Random effects for the linear and quadratic slopes for patients were included.

Unadjusted model-based trajectories were graphed, stratified by patient characteristics.

In multivariable models, interaction terms between fixed effect linear time and each covariate were used to test whether the rate of change varied significantly according to values of each covariate, after accounting for non-linear temporal patterns and individual trajectories. Models were used to estimate the mean difference in percentage weight change associated with each covariate at specific time points, namely 6 and 12 months. P-values for the interaction fixed effects were calculated based on the t-distribution. Each covariate was first evaluated separately, and resulting p-values were adjusted for multiple comparisons using the false discovery rate (FDR) method (Benjamini-Hochberg procedure).^25^ Covariates with FDR-adjusted *p*<0.05 were subsequently included in an initial full multivariable model. To build a parsimonious model, backward selection was employed, such that only covariates with *p*<0.05 (considered statistically significant) in the multivariable model (not FDR-adjusted) were retained.

Laboratory-based measurements with missing data were each evaluated in a separate multivariable model among patients with available data. Select covariates, including calendar time, health system, age, sex, pretreatment weight, and dose, were retained irrespective of p-value. Analyses were performed using R software, version 4.2.3.

## Results

### Cohort selection

Among 134,806 adults with linked EHR and Exome+^®^ sequencing data, 7,775 had a first-time semaglutide prescription documented between Mar 22, 2018 and Feb 23, 2024. Of these individuals, 1,923 met all inclusion criteria (see Methods and Figure 1). The most common reason for exclusion was having too short a duration of semaglutide use (n=3,789). The analytic cohort had a median follow-up duration of 9.4 months (IQR: 6.8-11.1) of on-treatment body weight measurements within 12 months. A median of 5 measurements (IQR: 3-8) were available per person.

**Figure 1:**
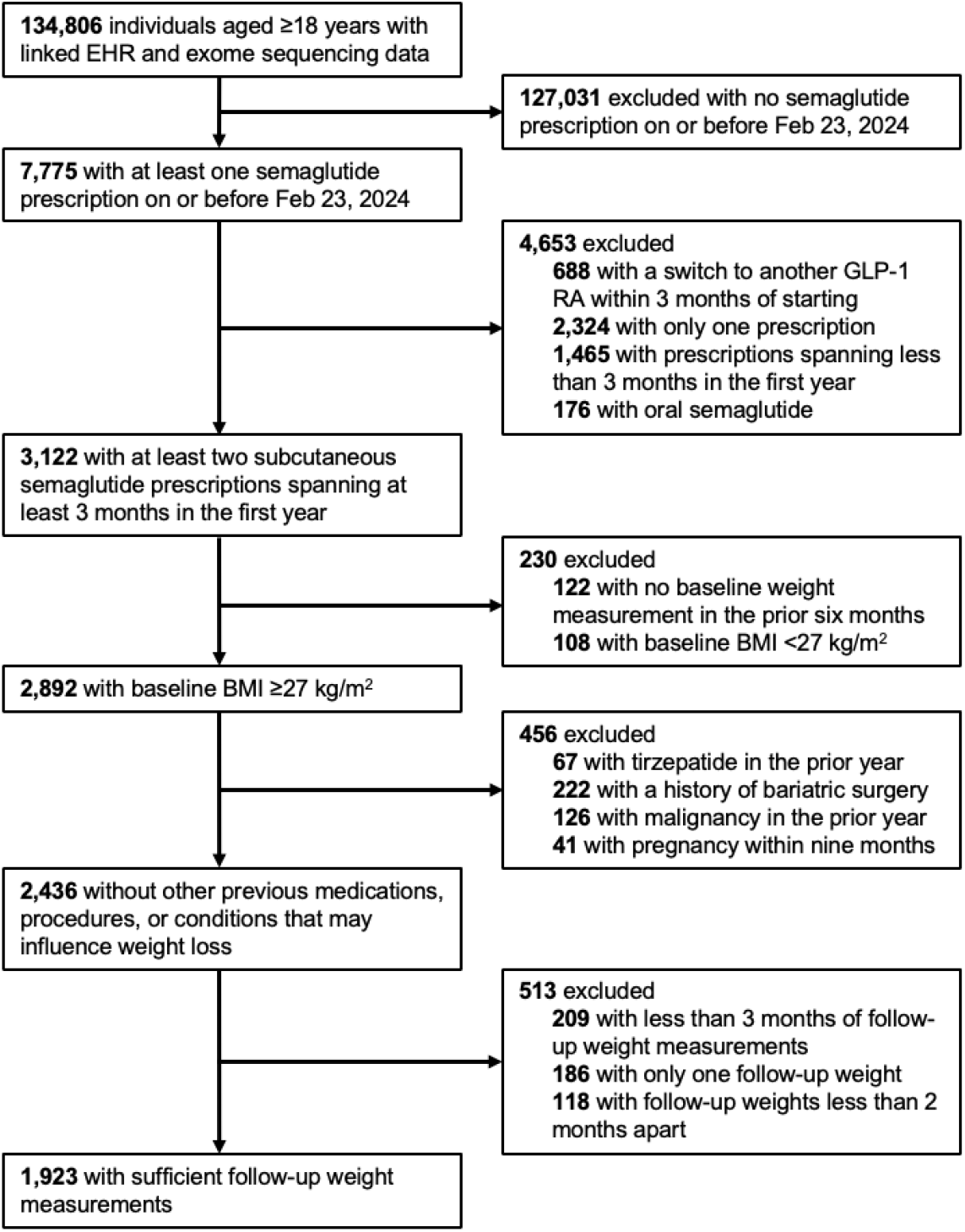
Cohort selection flowchart. GLP-1 RA=glucagon-like peptide 1 receptor agonist. EHR=electronic health record.

### Patient characteristics

Table 1 presents patient characteristics overall and stratified by the highest semaglutide dose reached. The recommended weekly maintenance dose for Ozempic (approved for T2D treatment) is 0.5 mg, adjustable to 1 or 2 mg as needed; for Wegovy (approved for weight loss), it is 2.4 mg, or 1.7 mg if the higher dose is not well-tolerated. In this real-world data, we observed maximum doses of 0.5 mg (n=772 [40.1%]), 1.0 mg (n=546 [28.4%]), 1.7 mg (n=82 [4.3%]), 2.0 mg (n=244 [12.7%]), and 2.4 mg (n=279 [14.5%]). The dose distribution stratified by T2D status is provided in the appendix (p 6).

**Table 1:**
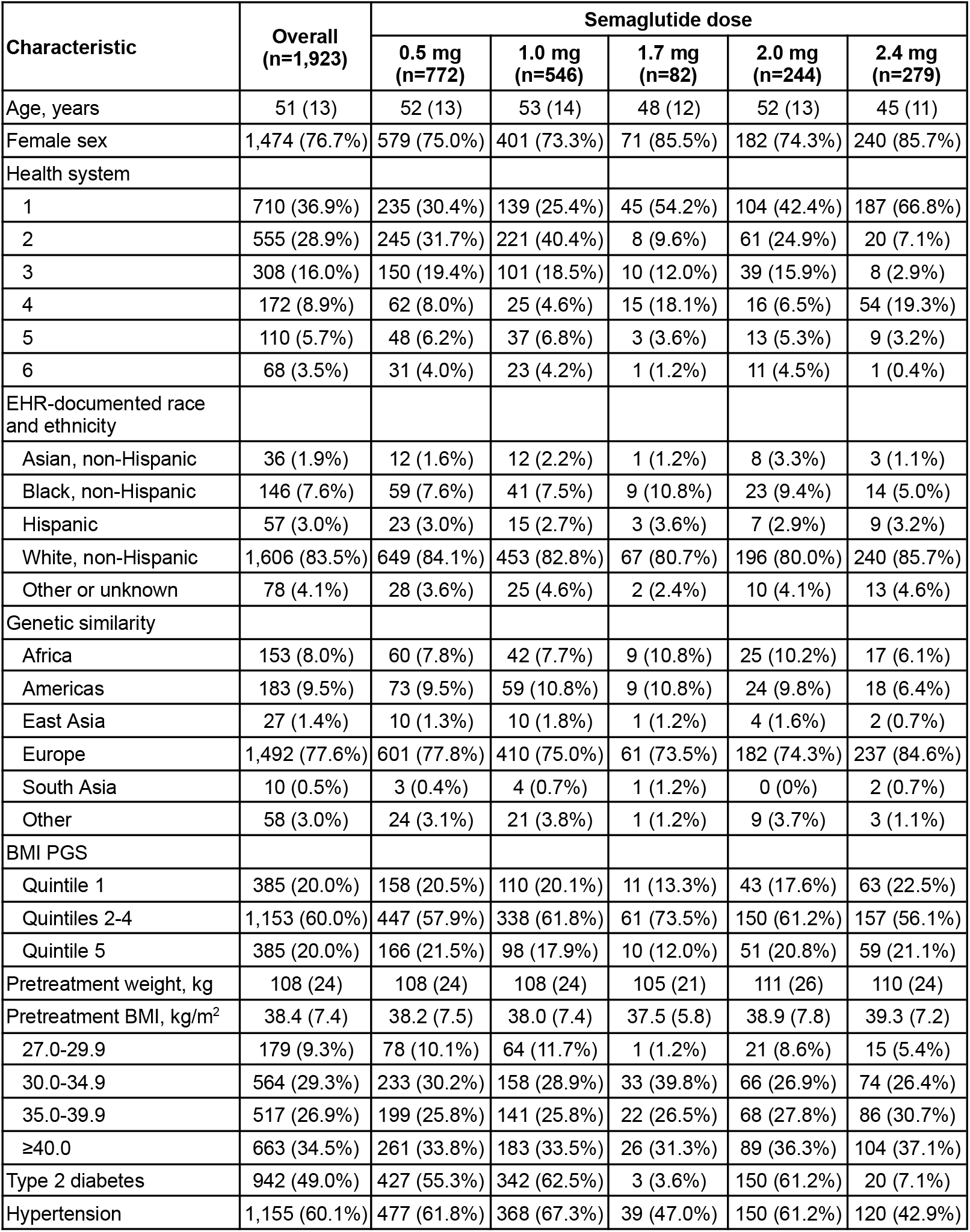
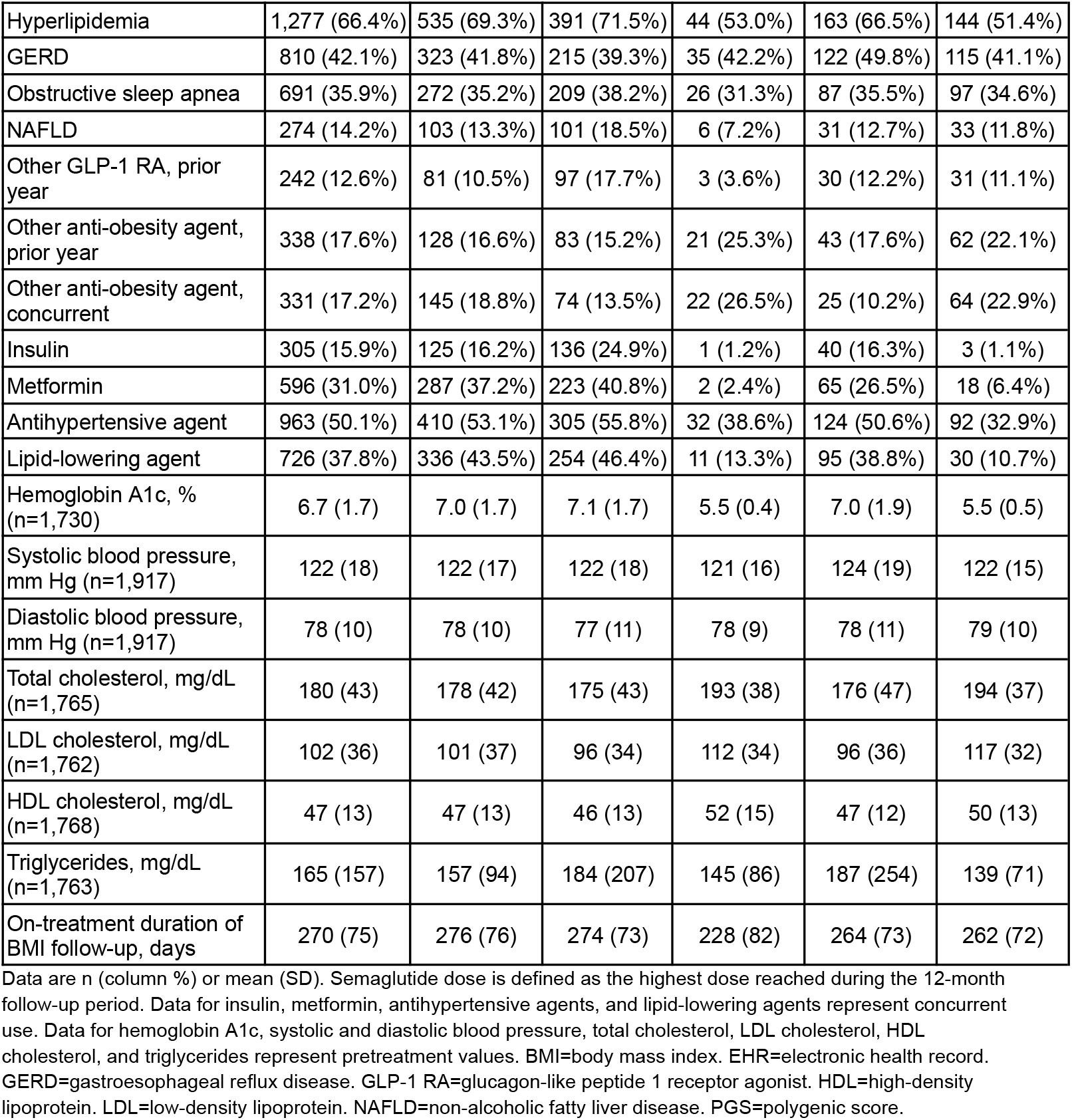
Patient characteristics.

| Characteristic | Overall<br>(n=1,923) | Semaglutide dose |  |  |  |  |
| --- | --- | --- | --- | --- | --- | --- |
|  |  | 0.5 mg<br>(n=772) | 1.0 mg<br>(n=546) | 1.7 mg<br>(n=82) | 2.0 mg<br>(n=244) | 2.4 mg<br>(n=279) |
| Age, years | 51 (13) | 52 (13) | 53 (14) | 48 (12) | 52 (13) | 45 (11) |
| Female sex | 1,474 (76.7%) | 579 (75.0%) | 401 (73.3%) | 71 (85.5%) | 182 (74.3%) | 240 (85.7%) |
| Health system |  |  |  |  |  |  |
| 1 | 710 (36.9%) | 235 (30.4%) | 139 (25.4%) | 45 (54.2%) | 104 (42.4%) | 187 (66.8%) |
| 2 | 555 (28.9%) | 245 (31.7%) | 221 (40.4%) | 8 (9.6%) | 61 (24.9%) | 20 (7.1%) |
| 3 | 308 (16.0%) | 150 (19.4%) | 101 (18.5%) | 10 (12.0%) | 39 (15.9%) | 8 (2.9%) |
| 4 | 172 (8.9%) | 62 (8.0%) | 25 (4.6%) | 15 (18.1%) | 16 (6.5%) | 54 (19.3%) |
| 5 | 110 (5.7%) | 48 (6.2%) | 37 (6.8%) | 3 (3.6%) | 13 (5.3%) | 9 (3.2%) |
| 6 | 68 (3.5%) | 31 (4.0%) | 23 (4.2%) | 1 (1.2%) | 11 (4.5%) | 1 (0.4%) |
| EHR-documented race and ethnicity |  |  |  |  |  |  |
| Asian, non-Hispanic | 36 (1.9%) | 12 (1.6%) | 12 (2.2%) | 1 (1.2%) | 8 (3.3%) | 3 (1.1%) |
| Black, non-Hispanic | 146 (7.6%) | 59 (7.6%) | 41 (7.5%) | 9 (10.8%) | 23 (9.4%) | 14 (5.0%) |
| Hispanic | 57 (3.0%) | 23 (3.0%) | 15 (2.7%) | 3 (3.6%) | 7 (2.9%) | 9 (3.2%) |
| White, non-Hispanic | 1,606 (83.5%) | 649 (84.1%) | 453 (82.8%) | 67 (80.7%) | 196 (80.0%) | 240 (85.7%) |
| Other or unknown | 78 (4.1%) | 28 (3.6%) | 25 (4.6%) | 2 (2.4%) | 10 (4.1%) | 13 (4.6%) |
| Genetic similarity |  |  |  |  |  |  |
| Africa | 153 (8.0%) | 60 (7.8%) | 42 (7.7%) | 9 (10.8%) | 25 (10.2%) | 17 (6.1%) |
| Americas | 183 (9.5%) | 73 (9.5%) | 59 (10.8%) | 9 (10.8%) | 24 (9.8%) | 18 (6.4%) |
| East Asia | 27 (1.4%) | 10 (1.3%) | 10 (1.8%) | 1 (1.2%) | 4 (1.6%) | 2 (0.7%) |
| Europe | 1,492 (77.6%) | 601 (77.8%) | 410 (75.0%) | 61 (73.5%) | 182 (74.3%) | 237 (84.6%) |
| South Asia | 10 (0.5%) | 3 (0.4%) | 4 (0.7%) | 1 (1.2%) | 0 (0%) | 2 (0.7%) |
| Other | 58 (3.0%) | 24 (3.1%) | 21 (3.8%) | 1 (1.2%) | 9 (3.7%) | 3 (1.1%) |
| BMI PGS |  |  |  |  |  |  |
| Quintile 1 | 385 (20.0%) | 158 (20.5%) | 110 (20.1%) | 11 (13.3%) | 43 (17.6%) | 63 (22.5%) |
| Quintiles 2-4 | 1,153 (60.0%) | 447 (57.9%) | 338 (61.8%) | 61 (73.5%) | 150 (61.2%) | 157 (56.1%) |
| Quintile 5 | 385 (20.0%) | 166 (21.5%) | 98 (17.9%) | 10 (12.0%) | 51 (20.8%) | 59 (21.1%) |
| Pretreatment weight, kg | 108 (24) | 108 (24) | 108 (24) | 105 (21) | 111 (26) | 110 (24) |
| Pretreatment BMI, kg/m<sup>2</sup> | 38.4 (7.4) | 38.2 (7.5) | 38.0 (7.4) | 37.5 (5.8) | 38.9 (7.8) | 39.3 (7.2) |
| 27.0-29.9 | 179 (9.3%) | 78 (10.1%) | 64 (11.7%) | 1 (1.2%) | 21 (8.6%) | 15 (5.4%) |
| 30.0-34.9 | 564 (29.3%) | 233 (30.2%) | 158 (28.9%) | 33 (39.8%) | 66 (26.9%) | 74 (26.4%) |
| 35.0-39.9 | 517 (26.9%) | 199 (25.8%) | 141 (25.8%) | 22 (26.5%) | 68 (27.8%) | 86 (30.7%) |
| ≥40.0 | 663 (34.5%) | 261 (33.8%) | 183 (33.5%) | 26 (31.3%) | 89 (36.3%) | 104 (37.1%) |
| Type 2 diabetes | 942 (49.0%) | 427 (55.3%) | 342 (62.5%) | 3 (3.6%) | 150 (61.2%) | 20 (7.1%) |
| Hypertension | 1,155 (60.1%) | 477 (61.8%) | 368 (67.3%) | 39 (47.0%) | 150 (61.2%) | 120 (42.9%) |
| Hyperlipidemia | 1,277 (66.4%) | 535 (69.3%) | 391 (71.5%) | 44 (53.0%) | 163 (66.5%) | 144 (51.4%) |
| GERD | 810 (42.1%) | 323 (41.8%) | 215 (39.3%) | 35 (42.2%) | 122 (49.8%) | 115 (41.1%) |
| Obstructive sleep apnea | 691 (35.9%) | 272 (35.2%) | 209 (38.2%) | 26 (31.3%) | 87 (35.5%) | 97 (34.6%) |
| NAFLD | 274 (14.2%) | 103 (13.3%) | 101 (18.5%) | 6 (7.2%) | 31 (12.7%) | 33 (11.8%) |
| Other GLP-1 RA, prior year | 242 (12.6%) | 81 (10.5%) | 97 (17.7%) | 3 (3.6%) | 30 (12.2%) | 31 (11.1%) |
| Other anti-obesity agent, prior year | 338 (17.6%) | 128 (16.6%) | 83 (15.2%) | 21 (25.3%) | 43 (17.6%) | 62 (22.1%) |
| Other anti-obesity agent, concurrent | 331 (17.2%) | 145 (18.8%) | 74 (13.5%) | 22 (26.5%) | 25 (10.2%) | 64 (22.9%) |
| Insulin | 305 (15.9%) | 125 (16.2%) | 136 (24.9%) | 1 (1.2%) | 40 (16.3%) | 3 (1.1%) |
| Metformin | 596 (31.0%) | 287 (37.2%) | 223 (40.8%) | 2 (2.4%) | 65 (26.5%) | 18 (6.4%) |
| Antihypertensive agent | 963 (50.1%) | 410 (53.1%) | 305 (55.8%) | 32 (38.6%) | 124 (50.6%) | 92 (32.9%) |
| Lipid-lowering agent | 726 (37.8%) | 336 (43.5%) | 254 (46.4%) | 11 (13.3%) | 95 (38.8%) | 30 (10.7%) |
| Hemoglobin A1c, % (n=1,730) | 6.7 (1.7) | 7.0 (1.7) | 7.1 (1.7) | 5.5 (0.4) | 7.0 (1.9) | 5.5 (0.5) |
| Systolic blood pressure, mm Hg (n=1,917) | 122 (18) | 122 (17) | 122 (18) | 121 (16) | 124 (19) | 122 (15) |
| Diastolic blood pressure, mm Hg (n=1,917) | 78 (10) | 78 (10) | 77 (11) | 78 (9) | 78 (11) | 79 (10) |
| Total cholesterol, mg/dL (n=1,765) | 180 (43) | 178 (42) | 175 (43) | 193 (38) | 176 (47) | 194 (37) |
| LDL cholesterol, mg/dL (n=1,762) | 102 (36) | 101 (37) | 96 (34) | 112 (34) | 96 (36) | 117 (32) |
| HDL cholesterol, mg/dL (n=1,768) | 47 (13) | 47 (13) | 46 (13) | 52 (15) | 47 (12) | 50 (13) |
| Triglycerides, mg/dL (n=1,763) | 165 (157) | 157 (94) | 184 (207) | 145 (86) | 187 (254) | 139 (71) |
| On-treatment duration of BMI follow-up, days | 270 (75) | 276 (76) | 274 (73) | 228 (82) | 264 (73) | 262 (72) |

Data are n (column %) or mean (SD). Semaglutide dose is defined as the highest dose reached during the 12-month follow-up period. Data for insulin, metformin, antihypertensive agents, and lipid-lowering agents represent concurrent use. Data for hemoglobin A1c, systolic and diastolic blood pressure, total cholesterol, LDL cholesterol, HDL cholesterol, and triglycerides represent pretreatment values. BMI=body mass index. EHR=electronic health record. GERD=gastroesophageal reflux disease. GLP-1 RA=glucagon-like peptide 1 receptor agonist. HDL=high-density lipoprotein. LDL=low-density lipoprotein. NAFLD=non-alcoholic fatty liver disease. PGS=polygenic score.

The mean age was 51 years, and the mean pretreatment BMI was 38.4 kg/m^2^. Patients were predominantly female (n=1,474 [76.7%]) and genetically similar to Europeans (n=1,492 [77.6%]), which was largely consistent with the characteristics of the broader network (appendix, pp 7-8). There was a high prevalence of metabolic disorder diagnoses including T2D (n=942 [49.0%]), hypertension (n=1,155 [60.1%]), hyperlipidemia (n=1,277 [66.4%]), obstructive sleep apnea (n=691 [35.9%]), and NAFLD (n=274 [14.2%]). Approximately one-fourth of patients (n=541 [28.1%]) had used either a different GLP-1 RA (n=242 [12.6%]) or another anti-obesity agent (n=338 [17.6%]) in the prior year.

### Percentage weight loss

The overall mean on-treatment percentage reduction in body weight was 3.2% (95% CI: 3.0-3.4) after 3 months, 5.4% (95% CI: 5.1-5.7) after 6 months, and 7.0% (95% CI: 6.3-7.7) after 12 months (Figure 2A). By 6 months, 50.3% of patients (95% CI: 47.5-53.1) had lost ≥5% of their body weight, and 20.7% (95% CI: 18.5-23.1) had lost ≥10% (Figure 2B). By 12 months, 36.2% (95% CI: 32.1-40.5) had lost ≥10%, and 15.5% (95% CI: 12.5-18.9) had lost ≥15%.

**Figure 2:**
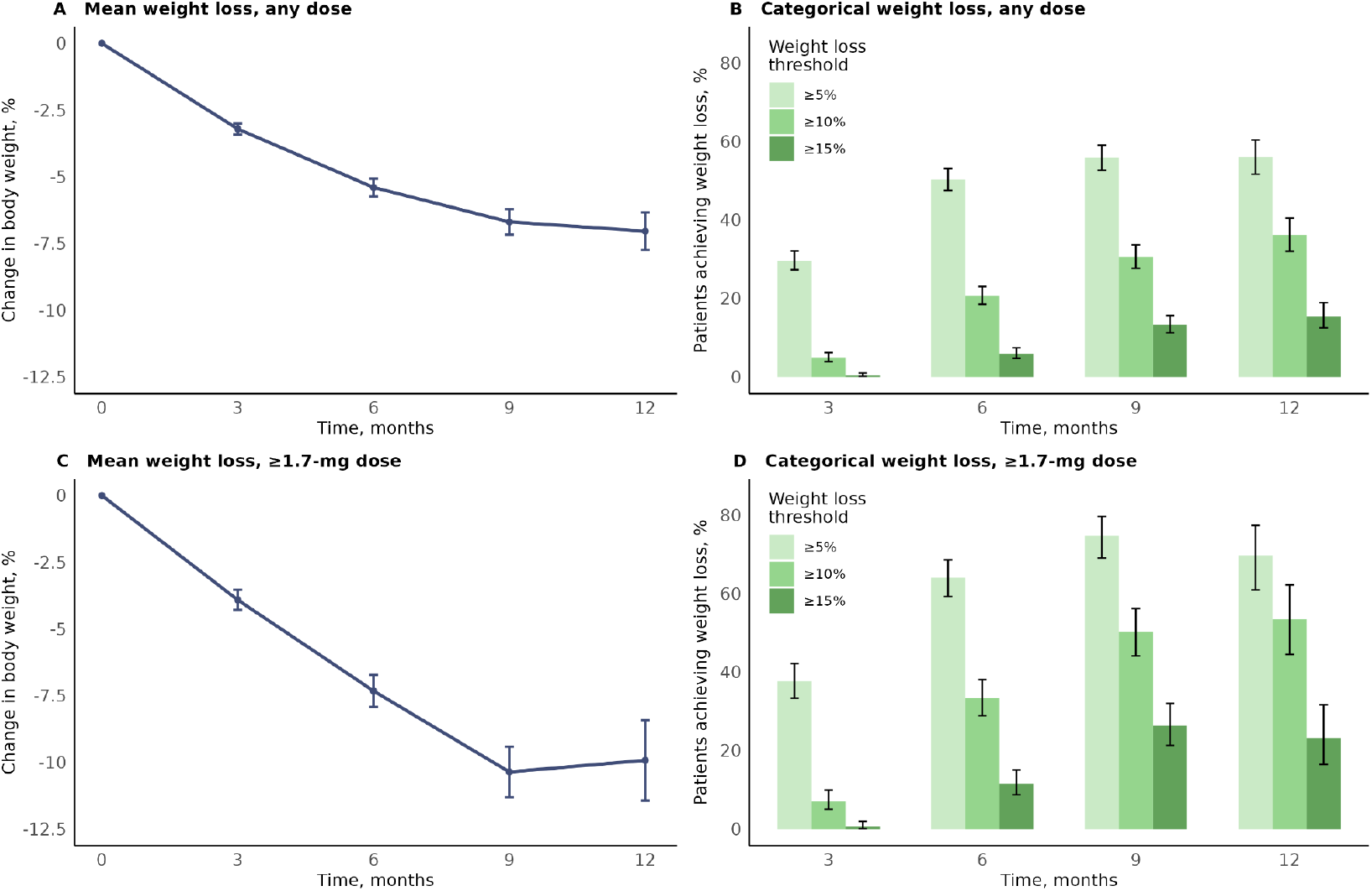
Percentage change in body weight at 3, 6, 9, and 12 months post-semaglutide initiation. The mean percentage change in body weight (panel A: any semaglutide dose; panel C: ≥1.7-mg semaglutide dose) and the proportions of patients achieving ≥5%,≥10%, and ≥15% weight loss (panel B: any semaglutide dose; panel D: ≥1.7-mg semaglutide dose) were computed at 3, 6, 9 and 12 months after starting semaglutide, among patients with an available weight measurement within a window of ±30 days. Error bars depict 95% confidence intervals.

Among patients who reached a semaglutide dose of at least 1.7 mg, percentage reductions at each time point were higher, with a mean reduction of 7.3% (95% CI: 6.7-7.9) after 6 months, and 9.9% (95% CI: 8.4-11.4) after 12 months (Figure 2C). By 6 months, 64.1% (95% CI: 59.3-68.6) had lost ≥5% of their body weight, and 33.3% (95% CI: 28.9-38.1) had lost ≥10% (Figure 2D). By 12 months, 53.5% (95% CI: 44.5-62.4) had lost ≥10%, and 23.3% (95% CI: 16.5-31.8) had lost ≥15%. Results for each dose are presented in the appendix (pp 9-10).

### Modeled weight loss trajectories and associated patient characteristics

Mixed effects models were built to identify patient characteristics associated with weight loss trajectories over the course of 12 months. Unadjusted modeled trajectories stratified by patient characteristics are displayed in Figure 3. Characteristics significantly associated with differential weight loss in the adjusted multivariable model are reported in Table 2 and described below.

**Table 2:**
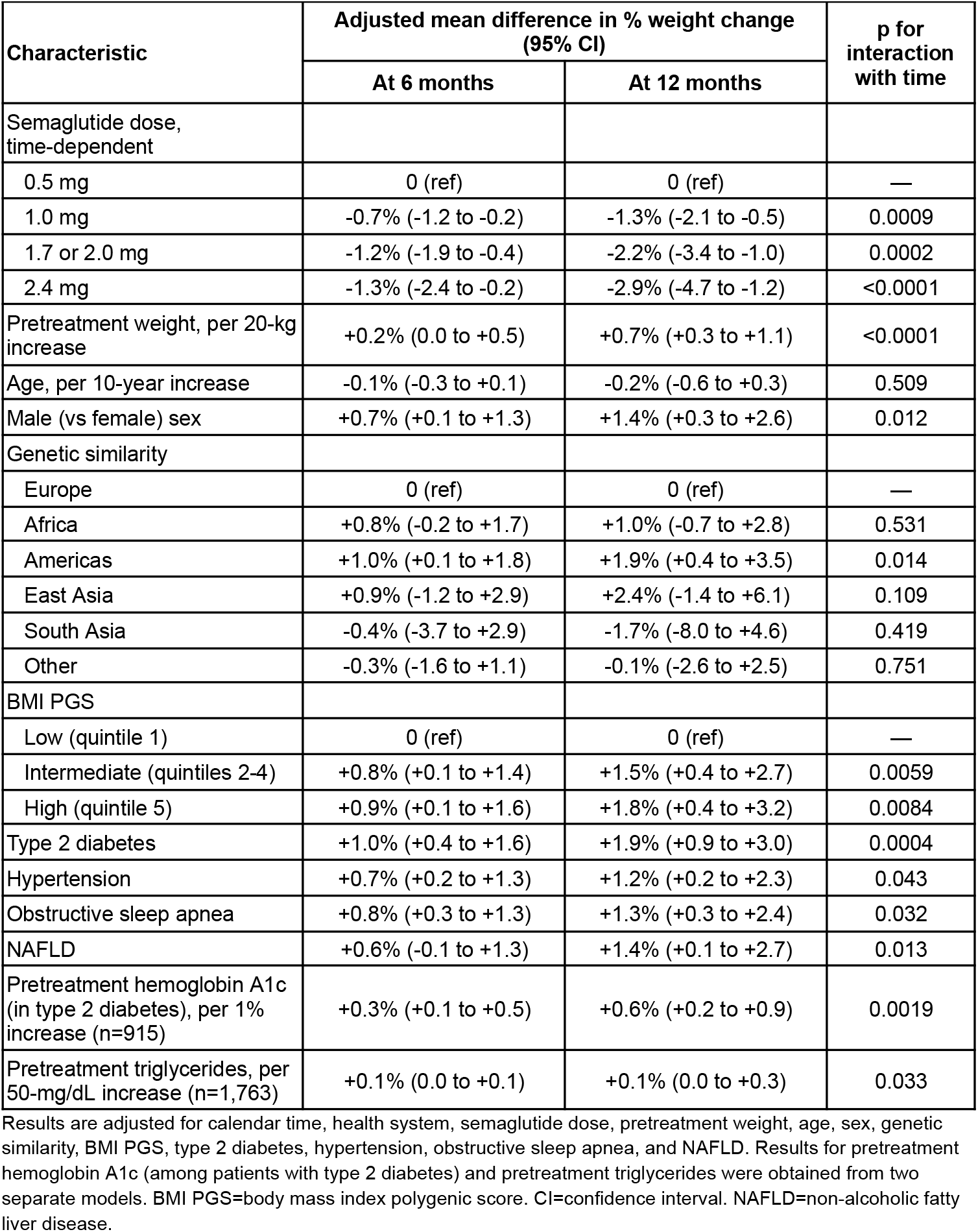
Adjusted mean differences in percentage weight change associated with patient characteristics.

**Figure 3:**
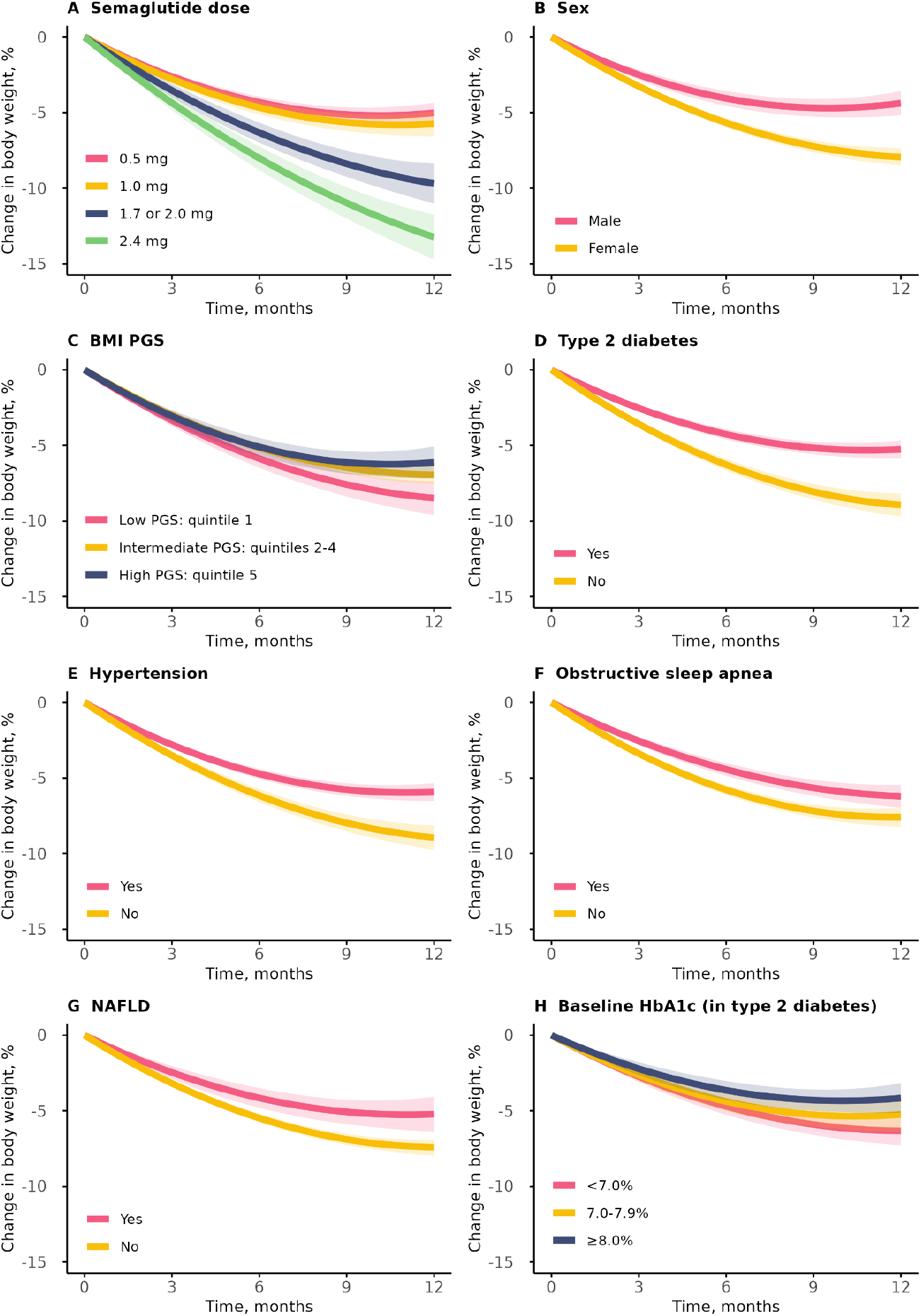
Modeled mean weight loss trajectories stratified by patient characteristics. Trajectories were modeled using linear mixed effects models with linear and quadratic terms for time, including random effects for the linear and quadratic slopes for individual patients. Shaded bands depict 95% confidence intervals. Semaglutide dose is defined as the highest dose reached during the 12-month period. BMI PGS=body mass index polygenic score. Hba1c=hemoglobin A1c. NAFLD=non-alcoholic fatty liver disease.

The full set of unadjusted and adjusted associations for all evaluated variables are provided in the appendix (pp 11-14).

#### Semaglutide dose

Compared to patients using a 0.5 mg semaglutide dose, those using 1.0 mg, 1.7 or 2.0 mg, or 2.4 mg doses lost, on average, an adjusted additional 1.3%, 2.2%, and 2.9% of their pretreatment body weight by 12 months, respectively (interaction with time, all p<0.001).

#### Demographic characteristics

The adjusted mean percentage weight loss among males was 1.4% lower after 12 months compared to females (p=0.012). This effect was stronger in the unadjusted model prior to controlling for pretreatment weight and other factors (3.4% less weight loss in males, p<0.0001). Significant adjusted differences among the participating health systems were detected. Older age was associated with less weight loss in the unadjusted analysis, but not the adjusted analysis.

#### Genetic factors

After 12 months, individuals with a low BMI PGS (bottom quintile) lost an adjusted average of 1.5% and 1.8% more body weight compared to those with intermediate (quintiles 2-4) or high (top quintile) BMI PGS, respectively (both p<0.01). PGS associations were similar when restricting the analysis to patients genetically similar to Europeans (appendix, p 15). Genetic similarity to those from the Americas was associated with an adjusted 1.9% lower weight loss compared to those genetically similar to Europeans (p=0.014), with no significant differences detected among other genetic similarity groups, either before or after controlling for other factors.

#### Metabolic disorders

Diagnoses including T2D, hypertension, obstructive sleep apnea, and NAFLD were each independently associated with an adjusted average of 1.2%-1.9% lower percentages of weight lost by 12 months (all p<0.05). In unadjusted analyses, hyperlipidemia, use of insulin, and other evaluated medications were also associated with less weight loss; however, these associations did not remain significant in the multivariable analysis.

#### Clinical and laboratory measurements

Greater pretreatment body weight was associated with less weight lost by 12 months (0.7% lower per 20-kg increase, p<0.0001). Each 50-mg/dL increase in pretreatment serum triglyceride levels was associated with an adjusted 0.1% reduction in weight lost by 12 months (p=0.033). Among individuals with T2D, each 1%-increase in pretreatment hemoglobin A1c was associated with an adjusted 0.6% lower weight loss (p=0.0019). Other serum lipid measurements were associated with weight loss in the unadjusted analysis, but not the multivariable analysis.

## Discussion

In this real-world clinico-genomic cohort study, we evaluated 12-month weight loss trajectories among semaglutide users with overweight or obesity, focusing on the influence of patient characteristics. To our knowledge, this is the first study to demonstrate that a lower genetic predisposition to obesity is associated with greater weight loss on semaglutide. In addition to confirming that male sex and T2D are independently linked to reduced weight loss, we detected novel associations with other metabolic disorders including hypertension, obstructive sleep apnea, and NAFLD. Our findings highlight the importance of advancing precision medicine in obesity management, underscoring the need for integrating genetic and metabolic risk profiling to improve weight loss expectations and enhance the personalization of GLP-1 RA therapies.

To comprehensively characterize real-world patterns, individuals with and without T2D were included, irrespective of whether they were prescribed Ozempic or Wegovy. Our data demonstrate a robust dose-response relationship, with users of 2.4-mg semaglutide (Wegovy) having lost an average of ∼13% of their weight, similar to what was observed in clinical trials.^1,2^ Ozempic has commonly been prescribed off-label,^26^ resulting in many patients not receiving the high 2.4-mg dose recommended for weight loss. In our cohort, only 44% of patients without T2D reached a dose ≥1.7 mg, and just 26% reached 2.4 mg. Other research suggests that non-adherence to the recommended dose escalation schedule is common, resulting in many users remaining on lower doses.^27^ Additionally, many patients discontinue quickly.^28^ In our study, 33% of individuals were excluded for having only a single prescription record. Possible reasons may include concerns about tolerability or side effects, costs, supply issues, or patient decisions related to lack of effectiveness or achievement of desired weight loss at lower doses.

This study uniquely incorporates genetic data through a PGS for BMI and identifies that a low PGS is associated with greater weight loss. The implication is that individuals with a high BMI despite a low genetic predisposition to obesity may experience a greater weight loss response to semaglutide, perhaps related to differences in the underlying behavioral and metabolic causes of obesity. This key finding persists after controlling for pretreatment weight, dose, and other factors, and highlights the potential of genetic risk stratification to optimize treatment outcomes. For many drugs, there are established associations between individual genetic variants and medication efficacy or adverse events, with well-documented pharmacogenetic recommendations.^29^ For semaglutide, a recent genome-wide association study investigated hemoglobin A1c reductions and weight loss in response to semaglutide in adults with T2D.^30^ However, to date, no genetic associations have been identified for weight loss with GLP-1 RAs. Here, the use of a PGS^23^ resulted in improved power over the ability to detect associations one variant at a time.

T2D was associated with less weight loss, which has been observed in other real-world studies^7,8,10^ and clinical trials.^1,2^ One possible reason is that the improved glucose control associated with semaglutide treatment reduces glycosuria, resulting in a positive calorie balance.^31^ We provide novel support for this hypothesis, as individuals with higher pretreatment serum hemoglobin A1c values–indicating poorer initial glucose control and greater potential for improvements in diabetes management–experienced less weight loss. Other possible reasons include altered microbiota, lower energy expenditure, and the use of concurrent diabetes medications (e.g., insulin).^31^ While insulin use was not significantly associated with weight changes in our adjusted analysis, it has previously been linked to weight gain.^32^

Hypertension, obstructive sleep apnea, NAFLD, and high serum triglyceride levels were also independently associated with less weight loss on semaglutide. These conditions are commonly comorbid with obesity and T2D, and involve metabolic, inflammatory, and hormonal dysregulations that may further hinder the drug’s effectiveness. Taken together, these findings underscore the substantial impact of metabolic health on semaglutide’s effectiveness. Further research is needed to identify the mechanisms linking metabolic comorbidities with GLP-1

### RA-associated weight loss patterns

Males lost significantly less weight than females, which aligns with sex differences reported elsewhere.^8,10^ One speculated reason is that differences in pretreatment body weight may lead to greater average drug exposure among females, thus, promoting more weight loss.^11^ Slightly more than half of the sex effect in our study was explained by differences in pretreatment weight and other measured factors. However, even after adjustment, a significant difference remained. Pharmacokinetic factors, such as differences in drug metabolism and serum concentration, and physiological differences, such as sex hormones and gastric emptying rates, may in part account for the observed disparity.^11^ Sex-specific factors should guide the clinical application of semaglutide for weight loss, with tailored dosing or additional support.

Our study was limited by the reliance on prescribing records to define intervals of semaglutide treatment and dosing patterns in the absence of information on how or whether medications were taken. There may be individuals included who did not take semaglutide or who did not comply with the recommended dosing schedule, or individuals with uninterrupted ongoing use who were classified as having discontinued (e.g., if they subsequently obtained prescriptions from a different health system). To minimize the impact of potential misclassification, we required that patients included have a record of multiple subcutaneous prescriptions from the same health system spanning at least a 3-month period in the first year. Although this reduced the sample size, it maximized the internal validity of analyses while maintaining the study’s advantage of representing semaglutide use in standard clinical practice, which can be more informative than clinical trials in some contexts. To that point, the associations detected could partially reflect differences in drug non-compliance. Furthermore, although a broad set of demographics, genetic factors, diagnoses, medications, and laboratory measurements were examined, residual and unmeasured confounding is possible. Specifically, data on behavioral factors (e.g., diet, exercise, sleep) and social factors (e.g., income, education) were not collected. The differences observed across some health systems in different geographic regions may in part reflect differences in such characteristics among the underlying patient populations.

Our study provides one of the most comprehensive assessments of the factors influencing semaglutide-associated weight loss in a real-world setting. Targeted treatment strategies can help maximize benefits for high responders and develop supportive measures for individuals with diminished responses. As the use of GLP-1 RAs continues to grow, these insights can inform both clinical practice and future research. Future studies should focus on identifying specific genetic variants and other factors that influence weight loss, side effect profiles, and additional health outcomes.

## Supporting information

Supplemental Data File: Concept IDs

## Data Availability

The Helix Research Network (HRN) data are available to qualified researchers upon reasonable request and with permission of the HRN Steering Committee and Helix. Researchers who would like to obtain the raw genotype data related to this study will be presented with a Data Use Agreement which requires that participants will not be reidentified and that no data will be shared between individuals, third parties, or uploaded onto public domains. The HRN encourages collaboration with scientific researchers on an individual basis. Examples of restrictions that will be considered in requests to access data include but are not limited to: 1) whether the request comes from an academic institution in good standing and will collaborate with our team to protect the privacy of the participants and the security of the data requested; 2) type and amount of data requested; 3) feasibility of the research suggested; and 4) amount of resource allocation for Helix and HRN member institutions required to support a collaboration.

## Article Information

### Contributors

M.E.L., N.T., K.M.S.B., A.B., and E.T.C. were responsible for the study concept and design. M.E.L. and E.T.C. were responsible for data analysis and for accessing and verifying the data. M.E.L. and E.T.C. were responsible for preparation of the first draft. D.S., C.N.C., C.A.A.C., D.P.J., D.A.O., J.J.G., N.L.W., W.L., and E.T.C. provided supervision over study activities. All authors provided support for acquisition and interpretation of the data. All authors provided critical revision of the manuscript for important intellectual content and gave approval of the submission of the manuscript for publication, with M.E.L. taking final responsibility for the decision to submit.

### Ethics declaration

The Helix Research Network cohorts were approved by the Salus IRB (reliance on Salus for all sites; approval number 21143), the WCG IRB (Western Institutional Review Board, WIRB-Copernicus Group; approval number 20224919), the MUSC Institutional Review Board for Human Research (approval number Pro00129083), and the University of Nevada, Reno Institutional Review Board (approval number 7701703417). All participants provided written informed consent prior to participation. All data used for research were de-identified.

## Acknowledgements

Funding was provided to the Desert Research Institute by the Renown Institute for Health Innovation and the Renown Health Foundation. Funding was provided to DRI by the Nevada Governor’s Office of Economic Development. Funding was provided to the myGenetics program by HealthPartners. We acknowledge the entire Helix bioinformatics and lab teams for their contributions to the production of the exome sequencing pipeline as well as the research administration team for coordinating the project. We thank all of the participants of DNA Answers, the Genetic Insights Project, the Healthy Nevada Project, In Our DNA SC, myGenetics, and The Gene Health Project.

