## Supplemental Appendix for "Influence of BMI-associated genetic variants and metabolic risk factors on weight loss with semaglutide: a longitudinal clinico-genomic cohort study"

### Semaglutide prescription records and associated data cleaning procedures

Appendix Figure 1 displays the recommended dose escalation schedules for Ozempic (labeled for type 2 diabetes management) and Wegovy (labeled for weight loss). All individuals initiating subcutaneous semaglutide are expected to start with 0.25 mg once-weekly for the first 4 weeks, followed by 0.5 mg once-weekly for the next 4 weeks. For weight loss (or if additional type 2 diabetes management is needed), additional dose escalations are recommended.

A Ozempic dosing schedule

B Wegovy dosing schedule

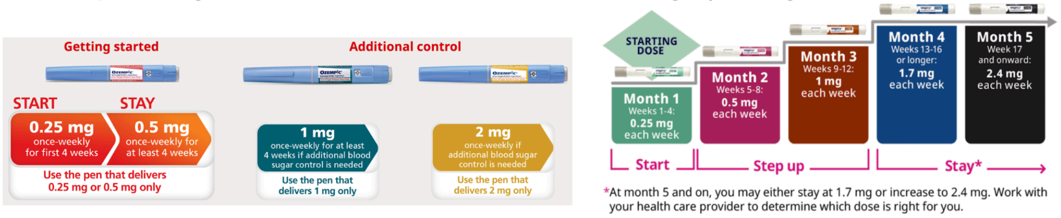


**Appendix Figure 1. Recommended dosing schedules for (A) Ozempic and (B) Wegovy.** Figure retrieved and adapted from https://www.ozempic.com and https://www.wegovy.com.

Appendix Table 1 displays the Observational Medical Outcomes Partnership (OMOP) Common Data Model (CDM) standard concepts representing subcutaneous semaglutide prescription records in our data. Each concept was associated with a specific brand name (Ozempic or Wegovy) and dose (0.25 mg, 0.5 mg, 1.0 mg, 1.7 mg, 2.0 mg, or 2.4 mg).

**Appendix Table 1. Subcutaneous semaglutide prescription records.**

| Drug concept ID | Drug concept name | Brand name | Dose |
|---|---|---|---|
| 1537596 | 0.5 ML semaglutide 0.5 MG/ML Auto-Injector | Wegovy | 0.25 mg |
| 1537597 | 0.5 ML semaglutide 0.5 MG/ML Auto-Injector [Wegovy] | Wegovy | 0.25 mg |
| 793147 | 0.25 MG, 0.5 MG Dose 1.5 ML semaglutide 1.34 MG/ML Pen Injector | Ozempic | 0.25 or 0.5 mg |
| 793152 | 0.25 MG, 0.5 MG Dose 1.5 ML semaglutide 1.34 MG/ML Pen Injector [Ozempic] | Ozempic | 0.25 or 0.5 mg |
| 741832 | 0.25 MG, 0.5 MG Dose 3 ML semaglutide 0.68 MG/ML Pen Injector | Ozempic | 0.25 or 0.5 mg |
| 741834 | 0.25 MG, 0.5 MG Dose 3 ML semaglutide 0.68 MG/ML Pen Injector [Ozempic] | Ozempic | 0.25 or 0.5 mg |
| 1537598 | 0.5 ML semaglutide 1 MG/ML Auto-Injector | Wegovy | 0.5 mg |
| 1537599 | 0.5 ML semaglutide 1 MG/ML Auto-Injector [Wegovy] | Wegovy | 0.5 mg |
| 37003617 | 3 ML semaglutide 1.34 MG/ML Pen Injector [Ozempic] | Ozempic | 1.0 mg |
| 37003616 | 3 ML semaglutide 1.34 MG/ML Pen Injector | Ozempic | 1.0 mg |
| 793154 | 1 MG Dose 1.5 ML semaglutide 1.34 MG/ML Pen Injector [Ozempic] | Ozempic | 1.0 mg |
| 793153 | 1 MG Dose 1.5 ML semaglutide 1.34 MG/ML Pen Injector | Ozempic | 1.0 mg |
| 1537600 | 0.5 ML semaglutide 2 MG/ML Auto-Injector | Wegovy | 1.0 mg |

| | | | |
|---|---|---|---|
| 1537601 | 0.5 ML semaglutide 2 MG/ML Auto-Injector [Wegovy] | Wegovy | 1.0 mg |
| 1537602 | 0.75 ML semaglutide 2.27 MG/ML Auto-Injector | Wegovy | 1.7 mg |
| 1537603 | 0.75 ML semaglutide 2.27 MG/ML Auto-Injector [Wegovy] | Wegovy | 1.7 mg |
| 780248 | 3 ML semaglutide 2.68 MG/ML Pen Injector | Ozempic | 2.0 mg |
| 780249 | 3 ML semaglutide 2.68 MG/ML Pen Injector [Ozempic] | Ozempic | 2.0 mg |
| 1537604 | 0.75 ML semaglutide 3.2 MG/ML Auto-Injector | Wegovy | 2.4 mg |
| 1537605 | 0.75 ML semaglutide 3.2 MG/ML Auto-Injector [Wegovy] | Wegovy | 2.4 mg |

For Ozempic, the same set of OMOP CDM concepts represents both 0.25 mg and 0.5 mg dosing; thus, it is not possible to distinguish between the two. Because all users are expected to escalate from 0.25 to 0.5 mg after 4 weeks, the two doses were grouped into one category for purposes of this analysis. Strictly speaking, the 0.5 mg dosing group described in this article represents either 0.25 or 0.5 mg. However, it is assumed that patients increased their dose in accordance with guidelines after 4 weeks, particularly since only patients with multiple prescriptions spanning at least 3 months were included.

We defined dosing intervals for each patient based on their chronology of prescriptions during the first 12 months of treatment. Recognizing that prescribing dates may not perfectly align with the actual dates of use, particularly given semaglutide's dose escalation schedules, we assumed that all patients used each dose for a minimum of 4 weeks (28 days) before escalating to the next dose in a stepwise manner (if prescribed on or prior to that date). Consequently, if patients received prescriptions for multiple doses on the same date, we assumed they adhered to the recommended dose escalation schedules before increasing from 0.5 to 1.0 mg (for both Ozempic and Wegovy), from 1.0 to 2.0 mg (for Ozempic), from 1.0 to 1.7 mg (for Wegovy), or from 1.7 to 2.4 mg (for Wegovy). If a patient subsequently had a prescription for a lower dose at a later date, it was assumed they stepped down in dose. Because stop dates were not reliably available, we assumed patients continued using semaglutide for 3 months after their final prescription, unless otherwise censored due to the end of available (or eligible) follow-up. In mixed effects models, semaglutide dose was treated as a time-dependent variable, using the current dose at the time of each weight measurement (following the above procedures).

**Data quality control procedures to ensure valid body weight measurements**

Within the framework of the OMOP CDM, weight measurements were identified using the standard concept ID 3025315 (body weight), which corresponds to LOINC code 29463-7 (body weight). Measurements recorded in units of oz or lbs were converted to kg. All patients included had a pretreatment weight measurement (defined as the most recent weight measurement on or before the semaglutide initiation date, within up to six months) and at least 2 follow-up weight measurements taken at least 2 months apart, spanning 3-12 months of treatment. To minimize the inclusion of implausible or invalid measurements, each follow-up measurement was compared against the respective patient's pretreatment measurement. In patients with at least a 12% increase or decrease in weight over the duration of follow-up, each measurement was evaluated for whether the estimated rate of change (compared to pretreatment weight) was greater than 12% per 3 months of follow-up (increase or decrease), based on the dates of measurements and assuming a consistent rate of change. To determine whether measurements were likely implausible, each flagged value was subsequently manually reviewed with respect to all earlier and later measurements for the respective patient. This procedure resulted in the exclusion of 18 weight measurements from this analysis.

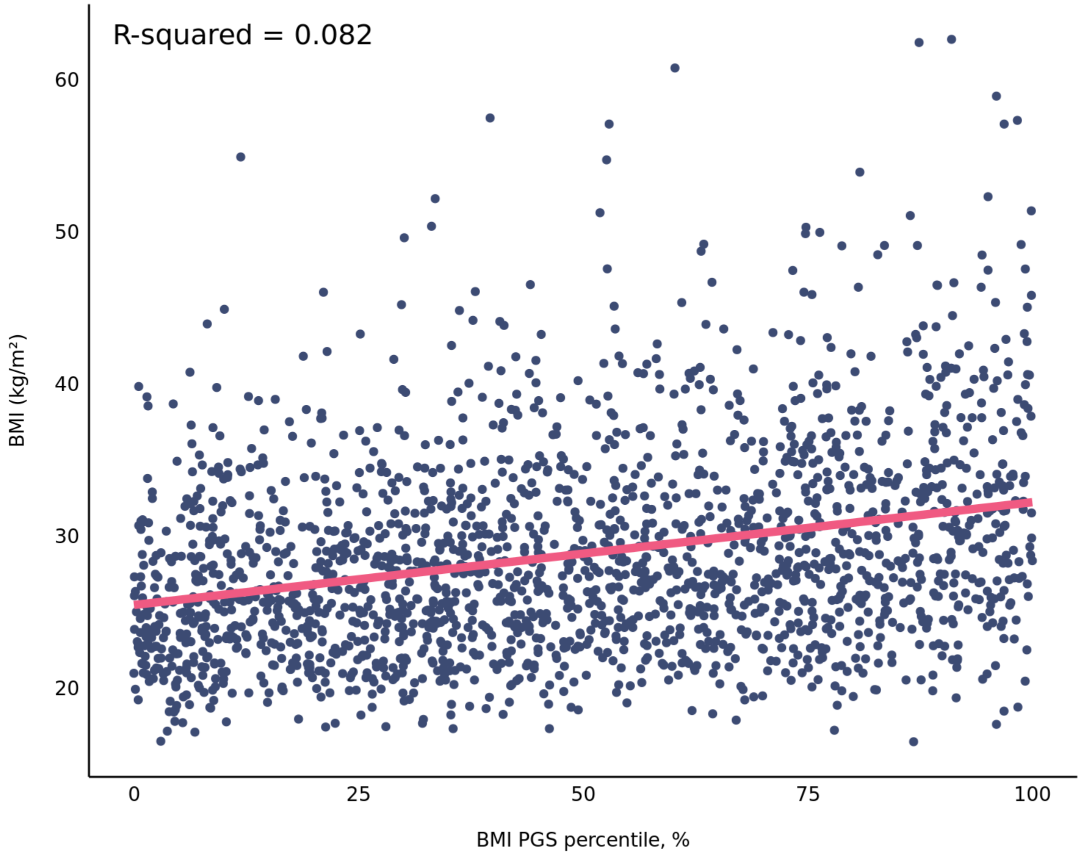


**Appendix Figure 2. Correlation between BMI PGS and the most recent BMI measurement among Helix Research Network participants with no history of GLP-1 RAs or other weight loss medications (n=96,914).** The scatterplot shows data points for 2,000 randomly selected patients. The regression line and R-squared is shown based on data from all 96,914 patients. R-squared values for each genetic similarity group are as follows: Africa: 0.0312; Americas: 0.0651; East Asia: 0.0846; Europe: 0.0894; South Asia: 0.0349; and Other: 0.0826. BMI=body mass index. GLP-1 RA=glucagon-like peptide 1 receptor agonist. PGS=polygenic score.

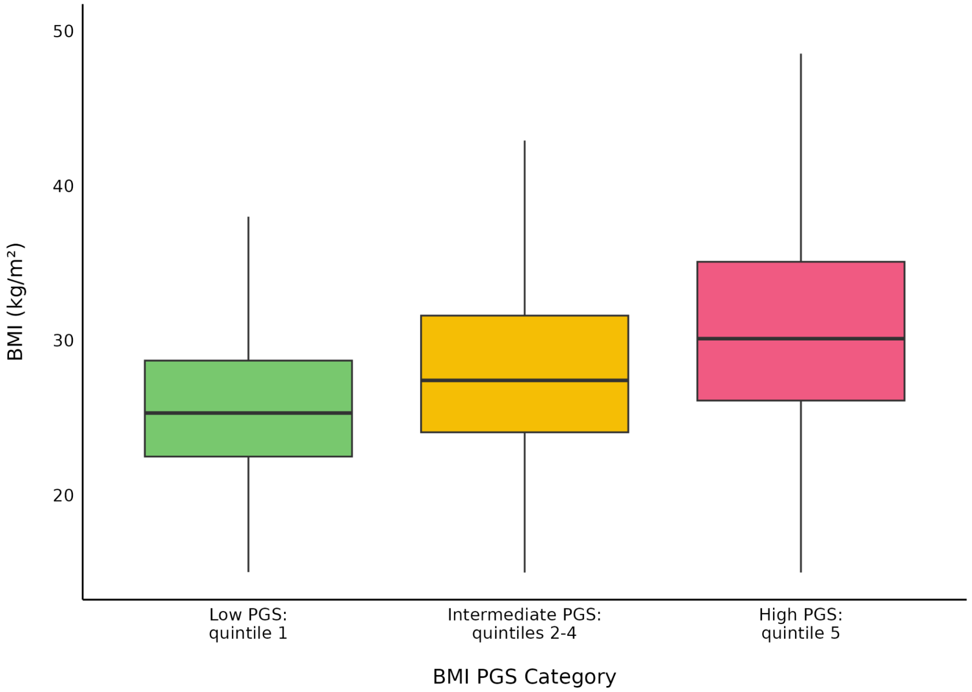


**Appendix Figure 3. Boxplot of most recent BMI stratified by BMI PGS category among Helix Research Network participants with no history of GLP-1 RAs or other weight loss medications (n=96,914).** The boxes show the 25th, 50th, and 75th percentiles, and the whiskers extend to the smallest and largest values within 1.5 times the interquartile range (IQR). The median BMI is as follows: low PGS: 25.3 kg/m$^2$ (IQR: 22.5-28.7); intermediate PGS: 27.5 kg/m$^2$ (IQR: 24.1-31.7); and high PGS: 30.3 kg/m$^2$ (IQR: 26.2-35.5) ($p<2.2e-16$). BMI=body mass index. GLP-1 RA=glucagon-like peptide 1 receptor agonist. PGS=polygenic score.

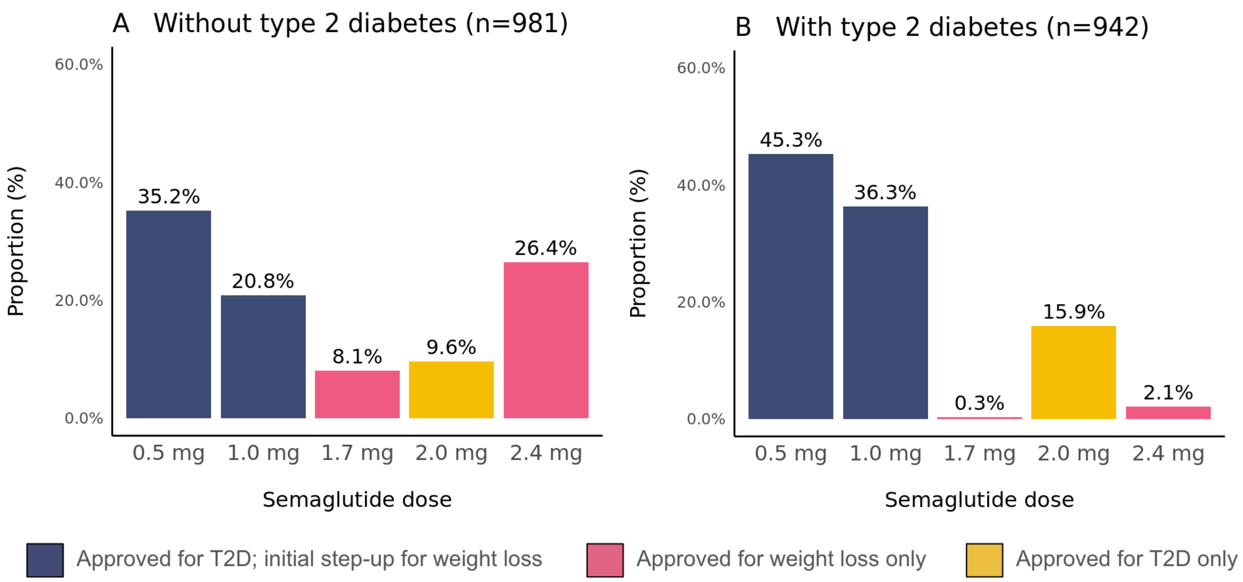


**Appendix Figure 4. Highest semaglutide dose reached in (A) patients without a type 2 diabetes diagnosis and (B) patients with a type 2 diabetes diagnosis.** T2D=type 2 diabetes.

**Appendix Table 2. Patient characteristics in the full Helix Research Network cohort compared to the semaglutide analysis cohort.**

| | Full Helix Research Network cohort (n=134,806) | Semaglutide analysis cohort (n=1,923) |
|---|---|---|
| Age, years | 52 (17) | 51 (13) |
| Female sex | 96,178 (71.3%) | 1,474 (76.7%) |
| Health system | | |
| 1 | 33,107 (24.6%) | 710 (36.9%) |
| 2 | 44,681 (33.1%) | 555 (28.9%) |
| 3 | 28,902 (21.4%) | 308 (16.0%) |
| 4 | 10,176 (7.5%) | 172 (8.9%) |
| 5 | 14,926 (11.1%) | 110 (5.7%) |
| 6 | 3,014 (2.2%) | 68 (3.5%) |
| EHR-documented race and ethnicity | | |
| Asian, non-Hispanic | 2,895 (2.1%) | 36 (1.9%) |
| Black, non-Hispanic | 5,447 (4.0%) | 146 (7.6%) |
| Hispanic | 3,417 (2.5%) | 57 (3.0%) |
| White, non-Hispanic | 115,594 (85.7%) | 1,606 (83.5%) |
| Other or unknown | 7,453 (5.5%) | 78 (4.1%) |
| Genetic similarity | | |
| Africa | 6,174 (4.6%) | 153 (8.0%) |
| Americas | 10,071 (7.5%) | 183 (9.5%) |
| East Asia | 2,746 (2.0%) | 27 (1.4%) |
| Europe | 111,199 (82.5%) | 1,492 (77.6%) |
| South Asia | 940 (0.7%) | 10 (0.5%) |
| Other | 3,676 (2.7%) | 58 (3.0%) |
| Pretreatment weight, kg | 84 (22) | 108 (24) |
| BMI, kg/m$^2$ | 29.7 (7.2) | 38.4 (7.4) |
| <27.0 | 51,080 (37.9%) | 0 (0%) |
| 27.0-29.9 | 22,734 (16.9%) | 179 (9.3%) |
| 30.0-34.9 | 25,893 (19.2%) | 564 (29.3%) |
| 35.0-39.9 | 13,734 (10.2%) | 517 (26.9%) |
| ≥40.0 | 10,987 (8.2%) | 663 (34.5%) |
| Type 2 diabetes | 15,417 (11.4%) | 942 (49.0%) |
| Hypertension | 44,797 (33.2%) | 1,155 (60.1%) |
| Hyperlipidemia | 59,488 (44.1%) | 1,277 (66.4%) |
| GERD | 36,033 (26.7%) | 810 (42.1%) |
| Obstructive sleep apnea | 18,507 (13.7%) | 691 (35.9%) |

| | | |
|---|---|---|
| NAFLD | 7,103 (5.3%) | 274 (14.2%) |

Data are n (column %) or mean (SD). BMI=body mass index. EHR=electronic health record. GERD=gastroesophageal reflux disease. NAFLD=non-alcoholic fatty liver disease.

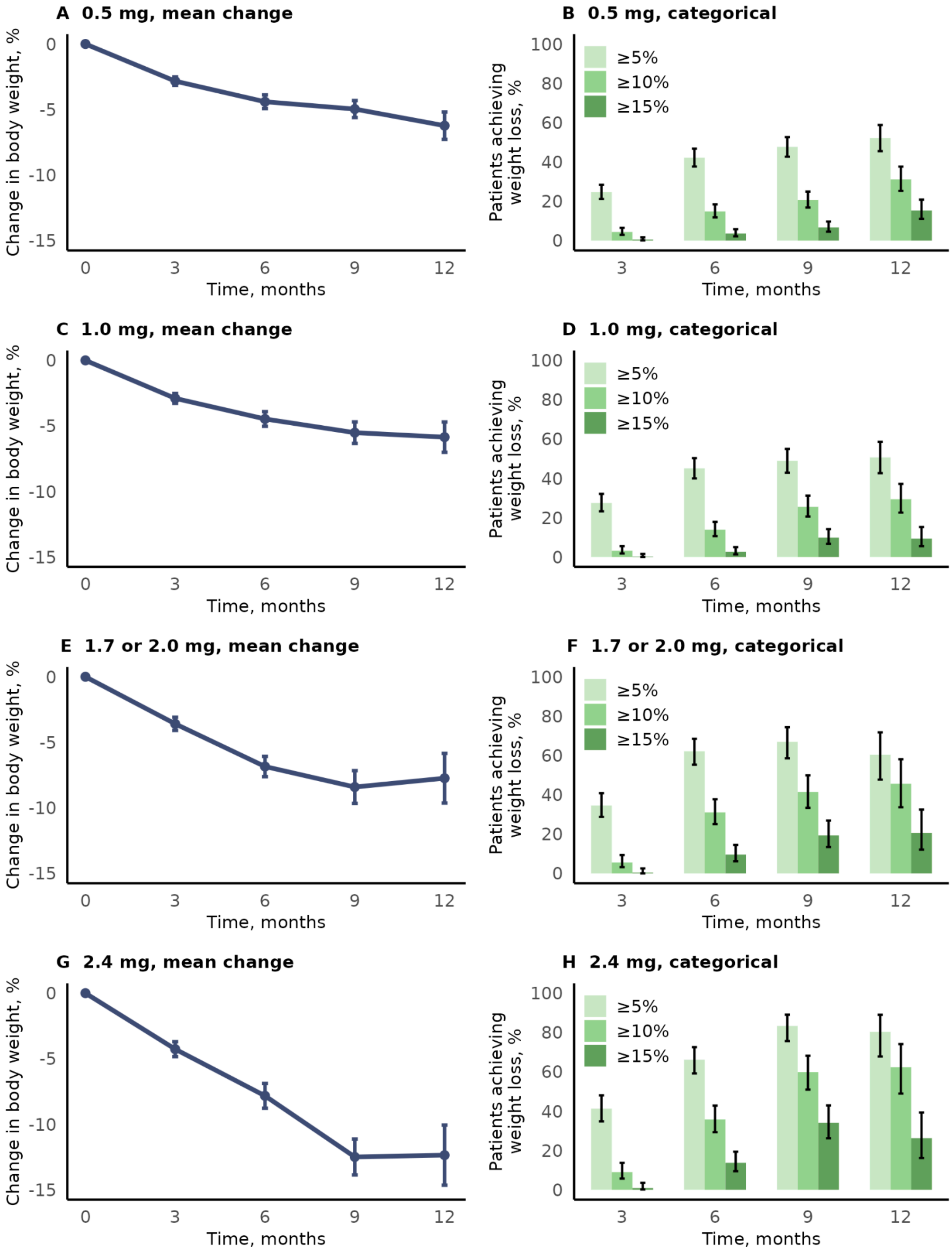


**Appendix Figure 5. Percentage change in body weight at 3, 6, 9, and 12 months post-semaglutide initiation, stratified by the highest semaglutide dose reached.** The mean percentage change in body weight (panels A, C, E, and G) and the proportions of patients achieving ≥5%, ≥10%, and ≥15% weight loss (panels B, D, F, and H) were computed at 3, 6, 9 and 12 months after starting semaglutide, among patients with an available weight measurement within a window of ±30 days. Error bars depict 95% confidence intervals.

**Appendix Table 3. Unadjusted mean differences in percentage weight change associated with patient characteristics.**

| | Unadjusted mean difference in % weight change (95% CI) | | FDR-adjusted p-value for interaction with time |
|---|---|---|---|
| | At 6 months | At 12 months | |
| Semaglutide dose, time-dependent | | | |
| 0.5 mg | 0 (ref) | 0 (ref) | — |
| 1.0 mg | -0.7% (-1.1 to -0.2) | -1.2% (-2.0 to -0.4) | 0.0086 |
| 1.7 or 2.0 mg | -1.3% (-2.0 to -0.5) | -2.4% (-3.6 to -1.2) | 0.0002 |
| 2.4 mg | -1.6% (-2.7 to -0.5) | -3.6% (-5.3 to -1.9) | <0.0001 |
| Pretreatment weight, per 20-kg increase | +0.4% (+0.2 to +0.6) | +1.0% (+0.6 to +1.4) | <0.0001 |
| Calendar time, per 1-year increase | -0.5% (-0.7 to -0.3) | -1.2% (-1.5 to -0.8) | <0.0001 |
| Age, per 10-year increase | +0.3% (+0.1 to +0.4) | +0.5% (+0.2 to +0.9) | 0.0098 |
| Male (vs female) sex | +1.6% (+1.0 to +2.2) | +3.4% (+2.3 to +4.5) | <0.0001 |
| Health system | | | |
| 1 | 0 (ref) | 0 (ref) | — |
| 2 | +1.7% (+1.1 to +2.3) | +3.7% (+2.5 to +4.9) | <0.0001 |
| 3 | +1.8% (+1.0 to +2.5) | +3.9% (+2.5 to +5.3) | <0.0001 |
| 4 | +0.8% (-0.1 to +1.7) | +1.2% (-0.5 to +2.9) | 0.398 |
| 5 | +1.0% (-0.2 to +2.1) | +2.2% (+0.1 to +4.3) | 0.033 |
| 6 | +0.5% (-0.9 to +1.8) | +1.2% (-1.4 to +3.8) | 0.34 |
| EHR-documented race and ethnicity | | | |
| White, non-Hispanic | 0 (ref) | 0 (ref) | — |
| Asian, non-Hispanic | -0.6% (-2.5 to +1.3) | -0.3% (-3.8 to +3.2) | 0.805 |
| Black, non-Hispanic | +1.0% (+0.1 to +2.0) | +1.6% (-0.2 to +3.4) | 0.254 |
| Hispanic | +1.4% (-0.1 to +2.8) | +2.4% (-0.4 to +5.3) | 0.168 |
| Other or unknown | +0.9% (-0.4 to +2.1) | +0.5% (-1.8 to +2.9) | 0.585 |
| Genetic similarity | | | |
| Europe | 0 (ref) | 0 (ref) | — |
| Africa | +0.9% (0.0 to +1.8) | +1.4% (-0.4 to +3.2) | 0.335 |
| Americas | +1.1% (+0.2 to +1.9) | +2.1% (+0.5 to +3.7) | 0.02 |
| East Asia | +0.8% (-1.3 to +2.9) | +2.0% (-1.9 to +6.0) | 0.254 |
| South Asia | -1.2% (-4.7 to +2.3) | -3.3% (-10.0 to +3.3) | 0.254 |
| Other | 0.0% (-1.5 to +1.4) | +0.4% (-2.3 to +3.2) | 0.537 |
| BMI PGS | | | |
| Low (quintile 1) | 0 (ref) | 0 (ref) | — |

| | | | |
|---|---|---|---|
| Intermediate (quintiles 2-4) | +0.7% (+0.1 to +1.4) | +1.5% (+0.3 to +2.7) | 0.017 |
| High (quintile 5) | +0.9% (+0.1 to +1.7) | +2.0% (+0.5 to +3.5) | 0.0059 |
| Type 2 diabetes | +1.9% (+1.4 to +2.4) | +3.8% (+2.9 to +4.8) | <0.0001 |
| Hypertension | +1.5% (+1.0 to +2.0) | +2.9% (+2.0 to +3.9) | <0.0001 |
| Hyperlipidemia | +1.1% (+0.5 to +1.6) | +1.8% (+0.8 to +2.8) | 0.0085 |
| GERD | +0.2% (-0.3 to +0.7) | +0.2% (-0.8 to +1.1) | 0.849 |
| Obstructive sleep apnea | +1.0% (+0.5 to +1.6) | +1.9% (+0.9 to +2.9) | 0.001 |
| NAFLD | +1.1% (+0.4 to +1.8) | +2.4% (+1.0 to +3.7) | 0.0004 |
| Other GLP-1 RA, prior year | +1.5% (+0.8 to +2.3) | +2.8% (+1.4 to +4.2) | 0.0008 |
| Other anti-obesity agent, prior year | -0.5% (-1.2 to +0.1) | -1.2% (-2.4 to +0.1) | 0.057 |
| Other anti-obesity agent, concurrent | -0.3% (-1.0 to +0.3) | -0.6% (-1.8 to +0.6) | 0.398 |
| Insulin | +2.0% (+1.4 to +2.7) | +3.8% (+2.6 to +5.1) | <0.0001 |
| Metformin | +1.5% (+0.9 to +2.0) | +3.2% (+2.2 to +4.2) | <0.0001 |
| Antihypertensive agent | +1.0% (+0.5 to +1.5) | +2.1% (+1.1 to +3.0) | <0.0001 |
| Lipid-lowering agent | +1.3% (+0.8 to +1.8) | +2.6% (+1.6 to +3.6) | <0.0001 |
| Pretreatment hemoglobin A1c (in type 2 diabetes), per 1% increase (n=915) | +0.4% (+0.2 to +0.5) | +0.7% (+0.4 to +1.1) | <0.0001 |
| Pretreatment hemoglobin A1c (in the absence of type 2 diabetes), per 1% increase (n=815) | +0.3% (-0.1 to +0.8) | +1.0% (+0.2 to +1.8) | 0.0026 |
| Systolic blood pressure, per 20-mm Hg increase (n=1,917) | -0.1% (-0.4 to +0.2) | 0.0% (-0.5 to +0.6) | 0.559 |
| Diastolic blood pressure, per 20-mm Hg increase (n=1,917) | -0.7% (-1.2 to -0.2) | -1.1% (-2.0 to -0.1) | 0.223 |
| Total cholesterol, per 20-mg/dL increase (n=1,765) | -0.1% (-0.2 to 0.0) | -0.3% (-0.5 to -0.1) | 0.012 |
| LDL cholesterol, per 20-mg/dL increase (n=1,762) | -0.2% (-0.4 to -0.1) | -0.5% (-0.8 to -0.2) | 0.0001 |
| HDL cholesterol, per 10-mg/dL increase (n=1,768) | -0.3% (-0.5 to -0.1) | -0.6% (-1.0 to -0.3) | 0.0008 |
| Triglycerides, per 50-mg/dL increase (n=1,763) | +0.1% (0.0 to +0.2) | +0.3% (+0.1 to +0.4) | 0.0004 |

BMI PGS=body mass index polygenic score. CI=confidence interval. EHR=electronic health record. FDR=false discovery rate. GERD=gastroesophageal reflux disease. GLP-1 RA=glucagon-like peptide 1 receptor agonist. HDL=high-density lipoprotein. LDL=low-density lipoprotein. NAFLD=non-alcoholic fatty liver disease.

**Appendix Table 4. Adjusted mean differences in percentage weight change associated with patient characteristics, for all variables evaluated in multivariable analysis.**

| | Adjusted mean difference in % weight change (95% CI) | | P-value for interaction with time |
|---|---|---|---|
| | At 6 months | At 12 months | |
| Semaglutide dose, time-dependent | | | |
| 0.5 mg | 0 (ref) | 0 (ref) | — |
| 1.0 mg | -0.7% (-1.2 to -0.2) | -1.3% (-2.1 to -0.5) | 0.0009 |
| 1.7 or 2.0 mg | -1.2% (-1.9 to -0.4) | -2.2% (-3.4 to -1.0) | 0.0002 |
| 2.4 mg | -1.3% (-2.4 to -0.2) | -2.9% (-4.7 to -1.2) | <0.0001 |
| Pretreatment weight, per 20-kg increase | +0.2% (0.0 to +0.5) | +0.7% (+0.3 to +1.1) | <0.0001 |
| Calendar time, per 1-year increase | -0.1% (-0.4 to +0.1) | -0.4% (-0.8 to +0.1) | 0.045 |
| Age, per 10-year increase | -0.1% (-0.3 to +0.1) | -0.2% (-0.6 to +0.3) | 0.509 |
| Male (vs female) sex | +0.7% (+0.1 to +1.3) | +1.4% (+0.3 to +2.6) | 0.012 |
| Health system | | | |
| 1 | 0 (ref) | 0 (ref) | — |
| 2 | +0.7% (+0.1 to +1.4) | +1.6% (+0.4 to +2.8) | 0.0021 |
| 3 | +1.6% (+0.9 to +2.4) | +3.7% (+2.2 to +5.1) | <0.0001 |
| 4 | +1.1% (+0.2 to +2.0) | +1.9% (+0.2 to +3.5) | 0.067 |
| 5 | +0.7% (-0.3 to +1.8) | +1.7% (-0.3 to +3.8) | 0.061 |
| 6 | +0.4% (-0.9 to +1.7) | +0.9% (-1.6 to +3.3) | 0.427 |
| Genetic similarity | | | |
| Europe | 0 (ref) | 0 (ref) | — |
| Africa | +0.8% (-0.2 to +1.7) | +1.0% (-0.7 to +2.8) | 0.531 |
| Americas | +1.0% (+0.1 to +1.8) | +1.9% (+0.4 to +3.5) | 0.014 |
| East Asia | +0.9% (-1.2 to +2.9) | +2.4% (-1.4 to +6.1) | 0.109 |
| South Asia | -0.4% (-3.7 to +2.9) | -1.7% (-8.0 to +4.6) | 0.419 |
| Other | -0.3% (-1.6 to +1.1) | -0.1% (-2.6 to +2.5) | 0.751 |
| BMI PGS | | | |
| Low (quintile 1) | 0 (ref) | 0 (ref) | — |
| Intermediate (quintiles 2-4) | +0.8% (+0.1 to +1.4) | +1.5% (+0.4 to +2.7) | 0.0059 |
| High (quintile 5) | +0.9% (+0.1 to +1.6) | +1.8% (+0.4 to +3.2) | 0.0084 |
| Type 2 diabetes | +1.0% (+0.4 to +1.6) | +1.9% (+0.9 to +3.0) | 0.0004 |
| Hypertension | +0.7% (+0.2 to +1.3) | +1.2% (+0.2 to +2.3) | 0.043 |
| Hyperlipidemia | +0.2% (-0.4 to +0.8) | 0.0% (-1.2 to +1.1) | 0.319 |
| Obstructive sleep apnea | +0.8% (+0.3 to +1.3) | +1.3% (+0.3 to +2.4) | 0.032 |
| NAFLD | +0.6% (-0.1 to +1.3) | +1.4% (+0.1 to +2.7) | 0.013 |

| | | | |
|---|---|---|---|
| Other GLP-1 RA, prior year | +0.7% (0.0 to +1.5) | +1.2% (-0.2 to +2.6) | 0.231 |
| Concurrent insulin | +0.9% (+0.2 to +1.6) | +1.3% (0.0 to +2.6) | 0.178 |
| Concurrent metformin | +0.3% (-0.3 to +0.9) | +0.7% (-0.4 to +1.8) | 0.153 |
| Concurrent antihypertensive agent | -0.1% (-0.7 to +0.4) | -0.2% (-1.2 to +0.8) | 0.844 |
| Concurrent lipid-lowering agent | +0.1% (-0.5 to +0.7) | -0.1% (-1.2 to +1.0) | 0.582 |
| Pretreatment hemoglobin A1c (in type 2 diabetes), per 1% increase (n=915) | +0.3% (+0.1 to +0.5) | +0.6% (+0.2 to +0.9) | 0.0019 |
| Pretreatment hemoglobin A1c (in the absence of type 2 diabetes), per 1% increase (n=815) | 0.0% (-0.5 to +0.4) | 0.0% (-0.8 to +0.9) | 0.668 |
| Total cholesterol, per 20-mg/dL increase (n=1,765) | +0.1% (-0.1 to +0.2) | +0.1% (-0.1 to +0.3) | 0.34 |
| LDL cholesterol, per 20-mg/dL increase (n=1,762) | 0.0% (-0.1 to +0.2) | 0.0% (-0.3 to +0.3) | 0.974 |
| HDL cholesterol, per 10-mg/dL increase (n=1,768) | 0.0% (-0.2 to +0.2) | +0.1% (-0.3 to +0.5) | 0.456 |
| Triglycerides, per 50-mg/dL increase (n=1,763) | +0.1% (0.0 to +0.1) | +0.1% (0.0 to +0.3) | 0.033 |

Adjusted for calendar time, health system, semaglutide dose, pretreatment weight, age, sex, genetic similarity, BMI PGS, type 2 diabetes, hypertension, obstructive sleep apnea, and NAFLD. BMI PGS=body mass index polygenic score. CI=confidence interval. EHR=electronic health record. GLP-1 RA=glucagon-like peptide 1 receptor agonist. HDL=high-density lipoprotein. LDL=low-density lipoprotein. NAFLD=non-alcoholic fatty liver disease.

**Appendix Table 5. Adjusted association between BMI PGS and percentage weight change only among patients genetically similar to Europeans.**

| Characteristic | Adjusted mean difference in % weight change (95% CI) | | p for interaction with time |
|---|---|---|---|
| | At 6 months | At 12 months | |
| BMI PGS | | | |
| Low (quintile 1) | 0 (ref) | 0 (ref) | — |
| Intermediate (quintiles 2-4) | +0.7% (-0.0 to +1.4) | +1.6% (+0.2 to +2.9) | 0.012 |
| High (quintile 5) | +0.9% (0.0 to +1.8) | +1.9% (+0.2 to +3.5) | 0.018 |

Adjusted for five European-specific principal components, calendar time, health system, semaglutide dose, pretreatment weight, age, sex, type 2 diabetes, hypertension, obstructive sleep apnea, and non-alcoholic fatty liver disease. BMI PGS=body mass index polygenic score. CI=confidence interval.
